# Efficacy and Safety of Beta Blockers for Patients with Myocardial Infarction in the Percutaneous Coronary Intervention Era: A Systematic Review and Meta-Analysis

**DOI:** 10.1101/2024.09.18.24313821

**Authors:** Ahmed Irtaza, Muhammad Junaid, Muhammad Saqlain, Ayesha Akram, Ayesha Khan, Areeba Shams Sarwari, Hussain Ali, Salman Ahsan, Tayyab Ahmed, Tuba Talib, Mohammad Ebad Ur Rehman, Huzaifa Ahmad Cheema, Adeel Ahmad, Wajeeh Ur Rehman, Abdul Wahab Arif, Sourbha S. Dani

## Abstract

**Background:** Beta-blockers are commonly prescribed to patients after MI. However, the evidence is based on studies conducted before the widespread use of PCI for treating MI. We conducted this meta-analysis to evaluate beta-blockers’ efficacy post-MI in the modern day and age.

**Methods:** We conducted our search using and one clinical trial registry to get RCTs and propensity-matched observational studies comparing the use of beta-blockers after MI to control in post-MI patients. The primary outcome of our analysis was the likelihood of all-cause mortality.

**Results:** Our analysis included 3 RCTs and 12 propensity-matched observational studies with a total study population of 102,794. Our results showed a statistically significant decrease in all-cause mortality in the beta-blocker group compared to the non-beta-blocker group (RR 0.63, 95% CI 0.47-0.83; p= 0.001). However, this decrease was not observed when only RCTs were considered (RR 0.91). Beta-blockers were also found to reduce the risk of MI and heart failure with RR of 0.86 (95% CI 0.75-1.00; p=0.05) and 0.84 (95% CI 0.73-0.97; p=0.02), respectively.

**Conclusion:** Beta-blockers effectively reduce mortality and decrease the risk of MI and heart failure without a significant increase in adverse effects. Thus, our findings support the contemporary use of beta blockers in post-MI patients. However, more long-term studies need to be done to determine the sustained benefits of beta blockers in the context of evolving cardiac care.

## Introduction

Beta-blockers have long been a cornerstone in managing acute myocardial infarction (AMI), primarily due to their established benefits in reducing mortality and morbidity observed in earlier clinical trials conducted before the advent of contemporary reperfusion strategies and secondary preventive therapies (1,2). These initial studies significantly influenced clinical guidelines and highlighted the efficacy of beta-blockers in patients with large infarcts and heart failure (HF) (3,4). However, the landscape of AMI treatment has dramatically evolved with the implementation of reperfusion therapies such as percutaneous coronary intervention (PCI) and the routine use of modern pharmacotherapies, including aspirin, statins, and ACE inhibitors/ARBs, prompting a re-evaluation of the role of beta-blockers today (5,6).

Recent meta-analyses and cohort studies have provided mixed evidence regarding beta-blockers’ efficacy in reducing all-cause mortality and cardiovascular events in contemporary AMI management(4,7). For instance, a systematic review of observational studies suggested a reduction in all-cause mortality with beta-blocker use, but this effect was significantly attenuated when accounting for publication bias and small-study effects(8). Furthermore, a comprehensive meta-analysis of randomized controlled trials (RCTs) stratified by era indicated a pronounced benefit in the pre-reperfusion era, with beta-blockers reducing mortality, recurrent myocardial infarction, and angina. However, these benefits appeared to diminish in the reperfusion era, with some adverse effects, such as increased heart failure and cardiogenic shock, becoming more prominent(9).

The ongoing debate about the necessity and duration of beta-blocker therapy post-AMI in the modern therapeutic landscape highlights the need for updated evidence(10). In contemporary practice, where PCI and modern pharmacotherapy are standard, beta-blockers’ role must be clearly defined to optimize patient outcomes without exposing them to unnecessary risks(6,11). The current guidelines still recommend beta-blockers post-AMI, but these recommendations are increasingly questioned as new evidence emerges(2,12).

Moreover, the specific patient characteristics and comorbid conditions that might influence beta-blockers’ effectiveness are not fully understood. For instance, beta-blockers might benefit patients with reduced ejection fraction or those with a history of heart failure. In contrast, their benefits in patients with preserved ejection fraction and no heart failure are less clear(4). Studies have shown that while beta-blockers reduce the risk of recurrent myocardial infarction and angina in the short term, they do not significantly impact long-term mortality in patients without heart failure(8,13).

In addition to efficacy concerns, the safety profile of beta-blockers is a topic of active investigation. While beta-blockers are generally well-tolerated, their side effects, such as bradycardia, hypotension, and fatigue, can impact patient adherence and quality of life. These side effects may be particularly problematic in older adults and those with comorbid conditions(14). The balance between the benefits and risks of beta-blocker therapy must be carefully considered, especially given the availability of other effective pharmacological agents.

To address these uncertainties, this meta-analysis evaluates the latest evidence on the post-discharge use of beta-blockers in patients with AMI in the current reperfusion era. By incorporating data from recent large-scale RCTs and propensity score-matched observational studies, we seek to provide a clearer understanding of the efficacy and safety of beta-blockers in this patient population, identify factors that may influence their effectiveness, and offer updated insights for clinical practice and future research.

## Methods

This meta-analysis was conducted following guidelines outlined in the Cochrane Handbook for Systematic Reviews of Interventions and described in compliance with the Preferred Reporting Items for Systematic Reviews and Meta-Analyses (PRISMA) statement(15). The protocol was registered with PROSPERO (CRD42024572013).

### Data Sources and Searches

A systematic literature search was undertaken on the Cochrane Library, MEDLINE, Embase, and ClinicalTrials.gov, which included studies published from January 2000 to June 2024. MeSH terms and keywords for “Myocardial Infarction” and “Adrenergic Beta-Antagonists” were used.

### Eligibility Criteria

The inclusion criteria included: (1) study design: RCTs and propensity-matched observational studies (2) patient population: patients with acute myocardial infarction (3) intervention: beta blockers after discharge; and (4) comparator: placebo/control.

The exclusion criteria consisted of (1) all study designs other than RCTs and observational studies that were propensity-matched, (2) studies conducted on patients not having acute MI, and (3) animal studies.

### Study Selection and Data Extraction

The studies identified by our search strategy were imported onto Rayyan (rayyan.ai), where duplicate articles were screened for and removed. Two authors (A.I. and A.S.) thoroughly inspected article titles, abstracts, and full texts of the remaining articles and studies, finalizing them following the pre-specified eligibility criteria. In the event of any disagreements concerning study selection, a senior investigator (M.R.) was consulted.

Data concerning study characteristics, including authors, study design, the origin of the study, patient population (including age and gender), interventions (including administration of beta-blockers or placebo), and primary and secondary outcomes were obtained. Data extraction was done independently by two authors (A.A. and A.K.) using a characterization table created using Microsoft Excel. A senior author was consulted in case of any disputes (M.R.).

### Outcomes

The primary outcome assessed was the risk of all-cause mortality. Secondary outcomes included the risk of cardiac death, MI, revascularization, MACE, heart failure, and stroke.

### Risk of Bias

We used the revised Cochrane Risk of Bias Tool for RCTs (RoB 2.0) to evaluate the risk of bias in the RCTs present in our analysis. RoB 2.0 assessed bias based on five domains: (1) bias arising from the randomization process; (2) bias due to deviations from intended interventions; (3) bias due to missing outcome data; (4) bias in the measurement of the outcome; and (5) bias resulting from the selection of the reported result(16). The Newcastle-Ottawa Scale (NOS) was used to assess the quality of the propensity-matched observational studies based on three aspects: the selection of study groups, the comparability of these groups, and the establishment of either the exposure or outcome of interest(17). Two investigators (M.S. and M.J.) assessed the risk of bias for each of the included studies as either high, low, or with some concerns of bias. Any disagreements regarding the risk of bias assessment were settled by a senior investigator (M.R.).

### Data Analysis

Meta-analyses were conducted using Review Manager (RevMan, Version 5.4; The Cochrane Collaboration, Copenhagen, Denmark). Risk ratios (RR) with 95% confidence intervals (CIs) for each study were extracted for all the dichotomous outcomes. The random-effects model with the Mantel-Haenszel method was used for the meta-analyses, and forest plots were constructed to display the results. The Higgins I^2^ statistic was calculated to evaluate statistical heterogeneity. We stratified all our analyses according to the study design (RCTs versus observational cohort studies).

## Results

### Search Results

The initial search retrieved a total of 6732 articles. After removing duplicates and screening titles and abstracts, 423 articles were eligible for full-text screening. Fifteen articles were included in the final systematic review and meta-analysis. Details of the search process and results are depicted in the PRISMA flowchart (Figure 1).

**Figure 1:**
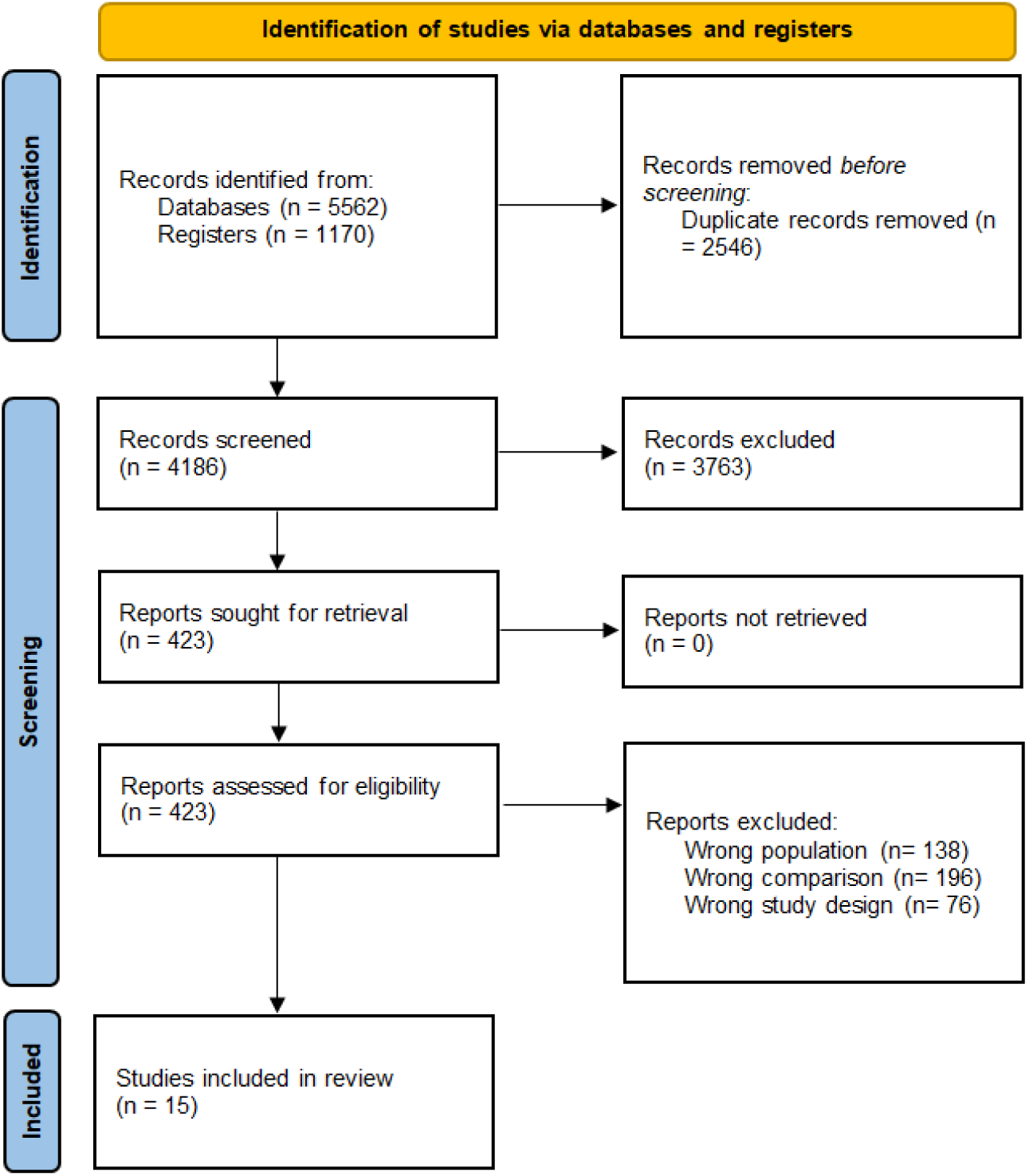
PRISMA Flowchart

### Study Characteristics

Our meta-analysis included 15 studies: 3 RCTs and 12 propensity-matched observational studies. Five studies were from South Korea (18–22), four from Japan (23), two from China (24,25), one from France(4), one from Sweden(26), one study included data from 7 Arabian Gulf countries namely Saudi Arabia, Kuwait, Bahrain, Qatar, UAE, Oman, and Yemen (27), and one had representative populations from Sweden, Estonia, and New Zealand (28). The years of publication ranged from 2013 to 2024. The total study population was 102794 patients, with 45301 (76 %) male. The median age in both groups, beta blocker and control, was 63.4 years. The characteristics of the included studies are summarized in Table 1.

**Table 1:**
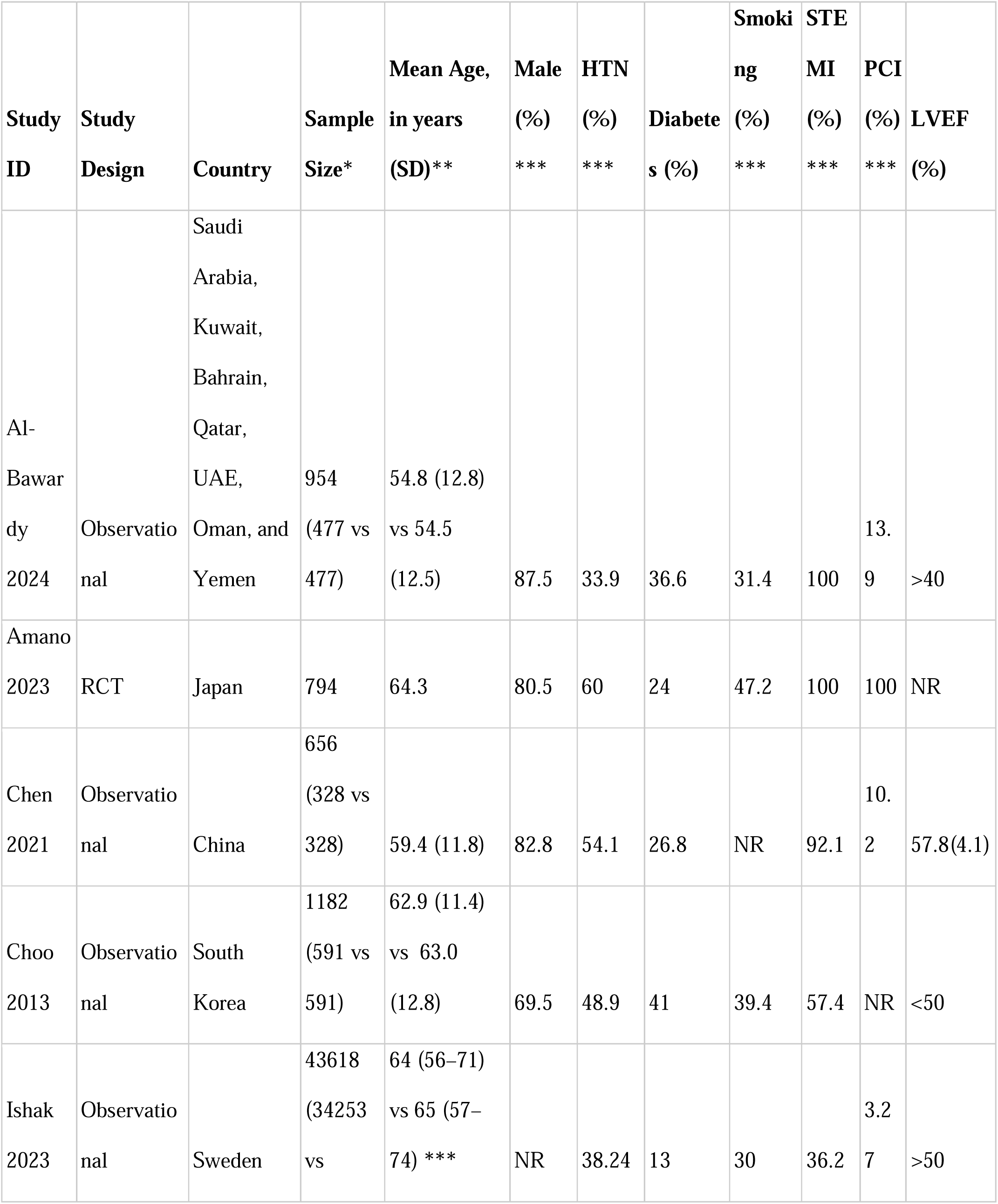
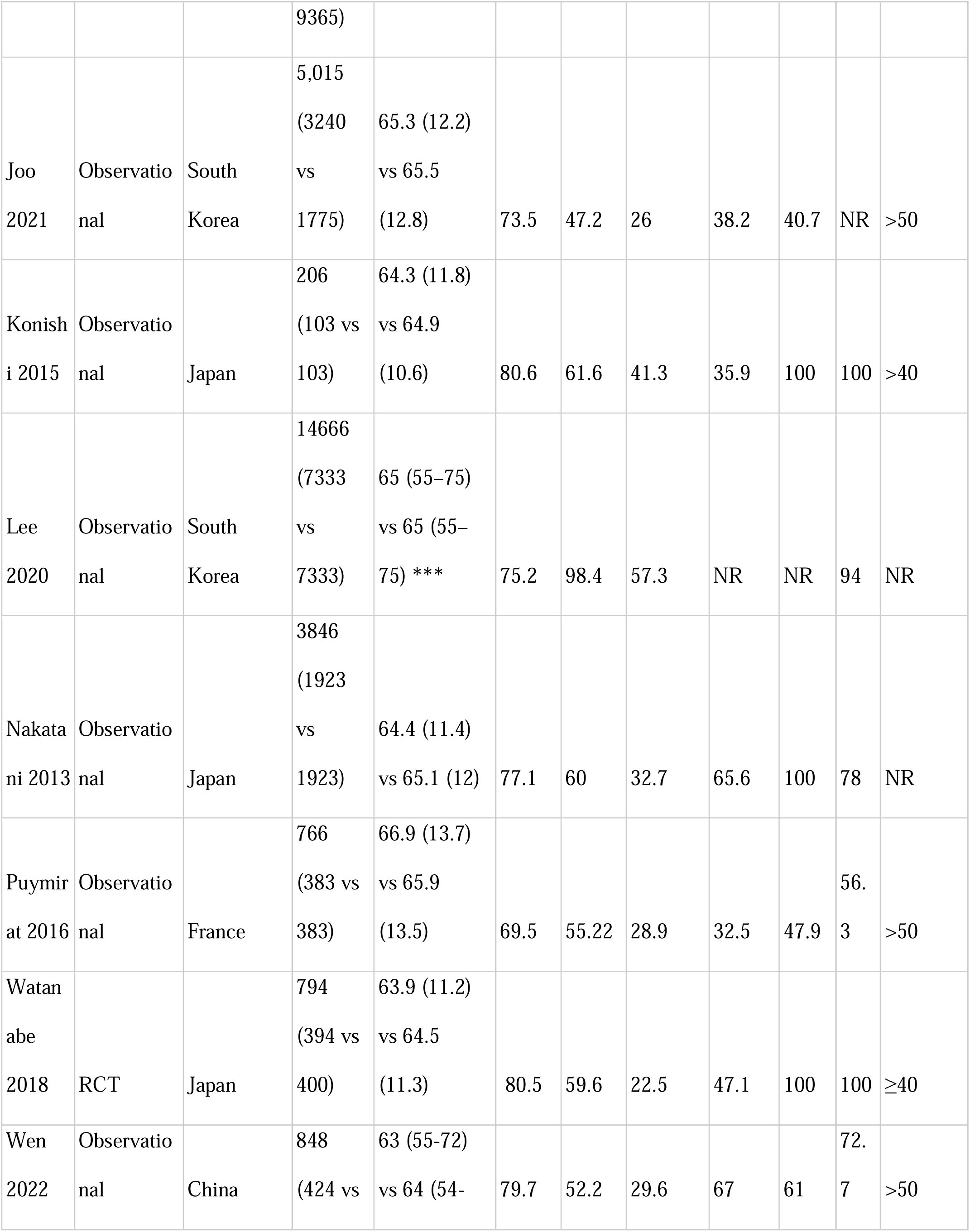
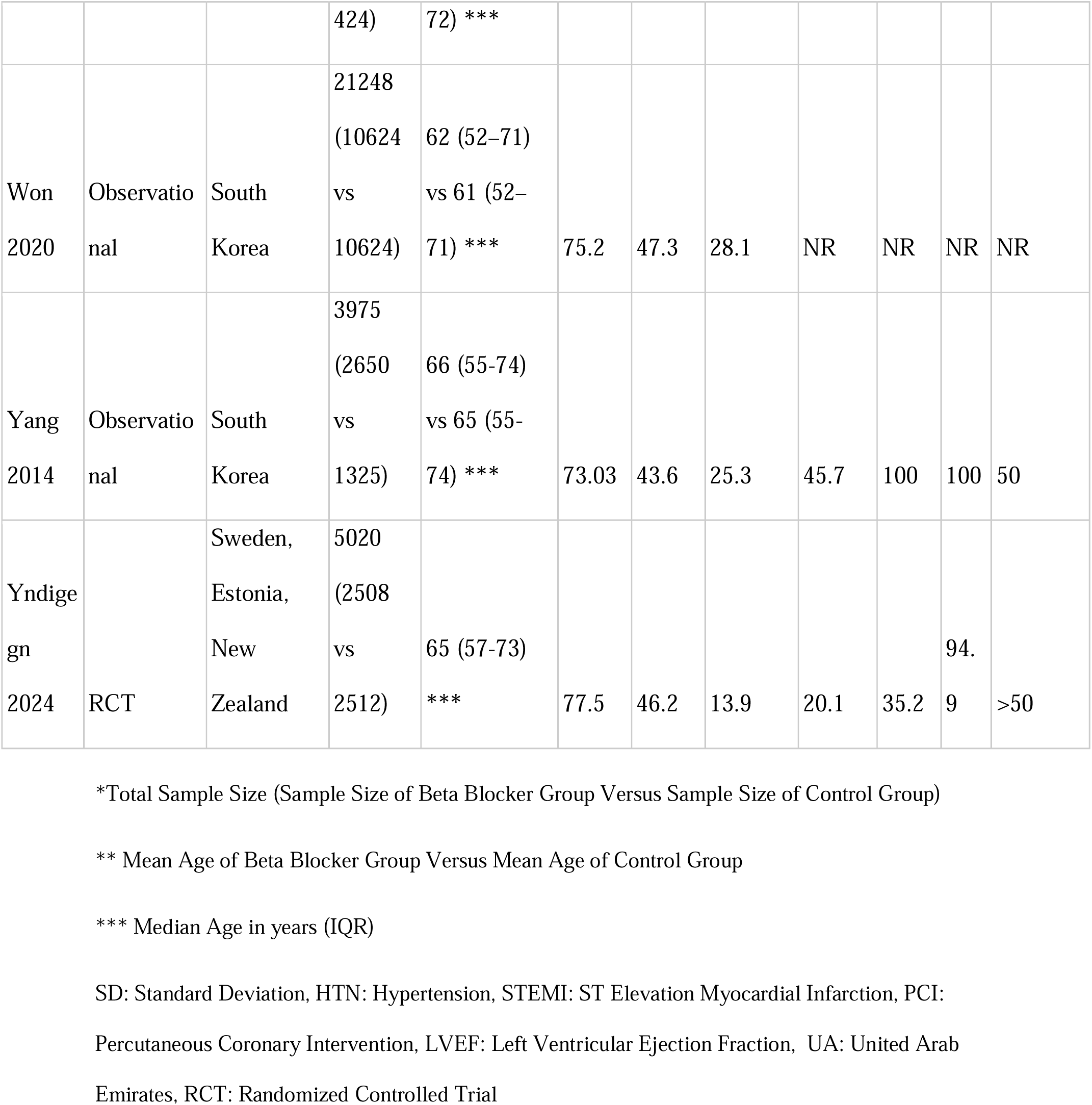
Characteristics of Included Studies.

### Risk of Bias in Included Studies

The risk of bias in included studies was assessed using Cochrane ROB 2.0 for RCTs and the Newcastle Ottawa Scale for observational studies. Of the three RCTs, Yndigegn et al. (28) showed a high concern for bias, particularly in the randomization domain. Amano et al. (29) and Watanbi et al. (30) showed some concern involving deviation from the intended intervention and measurement of the outcome domain. The Newcastle Ottawa scale score for observational studies ranged from 7-9, indicating a low risk of bias. The summary of risk of bias assessments is depicted in Supplementary Figure 1 and Supplementary Table 1.

### Meta-Analysis of Primary Outcome (All-Cause Mortality)

Fifteen studies were included in the analysis of all-cause mortality with 105,034 patients (67254 beta blockers vs 37780 control). There was a significant difference in all-cause mortality between the two groups, with a reduced risk of all-cause mortality in the beta-blockers group as compared to the control group (RR 0.63, 95% CI 0.47-0.83; p= 0.001; I^2^=96%) (Figure 2). In the RCT subgroup, beta-blockers did not significantly improve mortality (RR 0.91, 95% CI 0.72-1.14; p= 0.39; I^2^=0%), whereas in the observational studies subgroup, mortality was significantly lowered by beta-blocker use (RR 0.58, 95% CI 0.42-0.80; p= 0.001; I^2^=97%; p_interaction_=0.03).

**Figure 2:**
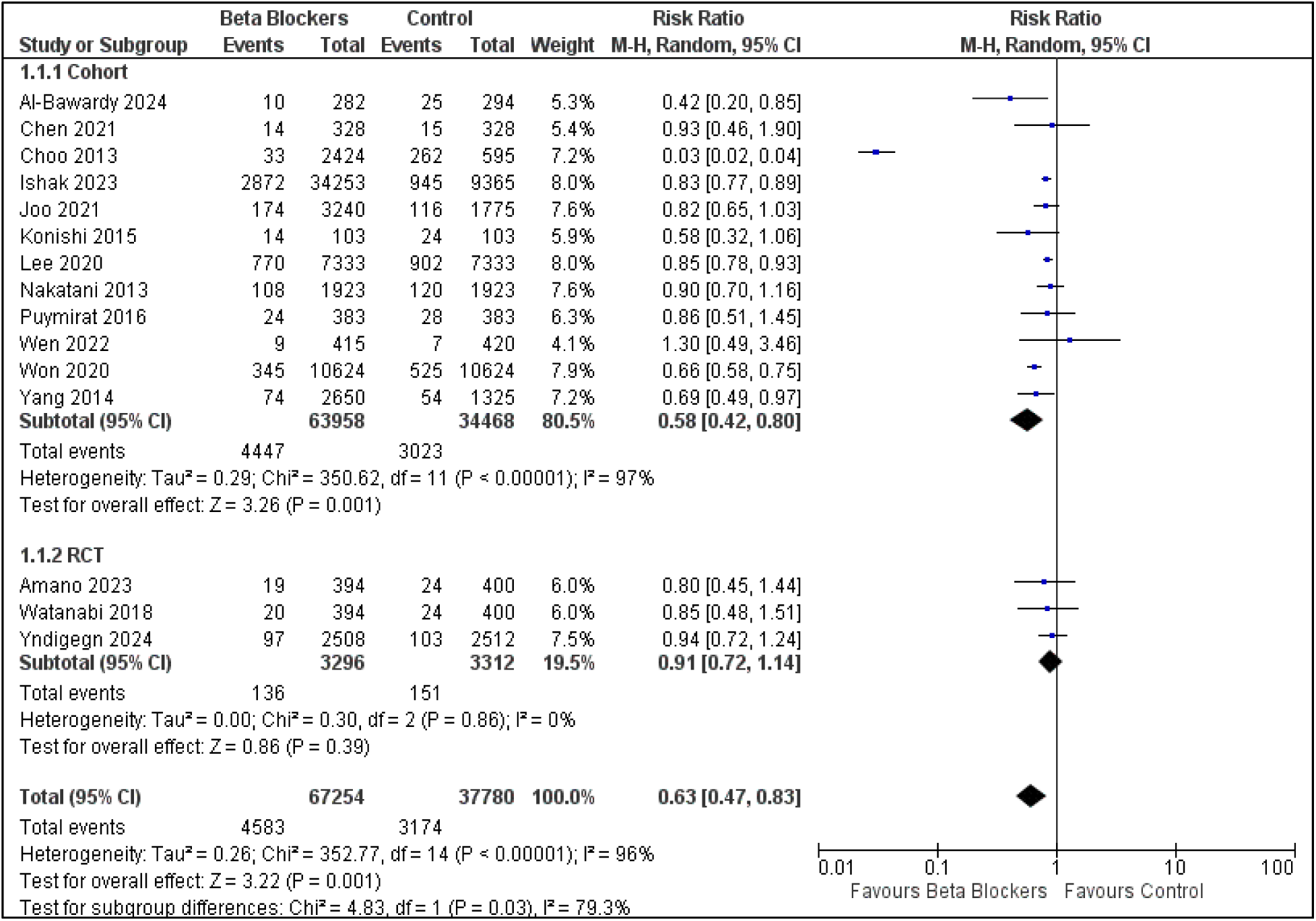
Forest Plot of All-Cause Mortality

### Meta-analysis of Secondary Outcomes

#### Cardiac Death

Eleven studies were included in the analysis for cardiac death with a total of 24926 patients (14762 beta-blockers vs 10164 control). No statistically significant association was found between beta-blocker use in MI patients and cardiac death (RR=0.56, 95% CI 0.25-1.25; p=0.16; I^2^=95%) (Supplementary Figure 7). There was no difference between the beta-blocker group and the control group in RCTs (RR 1.17, 95% CI 0.78-1.75; p= 0.45; I^2^=0%) and observational studies (RR 0.43, 95% CI 0.16-1.17; p= 0.10; I^2^=96%,p_interaction_=0.07).

#### Myocardial Infarction

Eight studies were included in the analysis of myocardial infarction with a total of 81299 patients (54478 beta-blockers vs 26821 control). A statistically significant difference was noted in the risk of myocardial infarction between the two groups, with reduced risk in the beta-blocker group as compared to the control group (RR 0.86, 95% CI 0.75-1.00; p=0.05; I^2^=41%) (Supplementary Figure 3). The results were comparable for RCTs and observational cohort studies (p_interaction_=0.6).

#### Major Adverse Cardiac Event (MACE)

Six studies were included in the analysis of MACE with a total of 12069 patients (7421 Beta Blockers vs 4648 Control). The analysis revealed no statistically significant difference in the risk of MACE between the two groups, with a p-value of 0.42. (RR 0.92, 95% CI 0.75-1.12; p=0.42; I^2^=65%) (Supplementary Figure 4). There was no difference between the results of RCTs and cohort studies (p_interaction_=0.78).

#### Stroke

Eight studies were included in the stroke analysis with a total of 80,040 patients (54197 Beta Blockers vs 25843 Control). The two regimens were comparable (RR 0.69, 95% CI 0.40-1.17; p=0.17; I^2^=93%) (Supplementary Figure 5). The results were similar between RCTs and observational studies (p_interaction_=0.22).

#### Heart Failure

Seven studies in our meta-analysis reported results for heart failure. The risk of heart failure in the beta-blocker group is lower than in the control group (RR=0.84, 95% CI 0.73-0.97; p=0.02; I^2^=0%) (Supplementary Figure 6). There was no difference in the results between RCTs and cohort studies (p_interaction_=0.18).

#### Revascularization

Six studies of our meta-analysis were included in the analysis for revascularization. No statistically significant association was found between beta-blocker use in MI patients and the need for revascularization (RR=0.97, 95% CI 0.90-1.04; p=0.43; I^2^=17%) (Supplementary Figure 7). The results were comparable for RCTs and observational studies (p_interaction_=0.57).

## Discussion

In this meta-analysis, including RCTs and propensity score-matched observational studies, we evaluated beta-blockers’ efficacy after discharge in patients with myocardial infarction. The use of beta-blockers after discharge was noted to be associated with a reduction in all-cause mortality, myocardial infarction, and heart failure. There were no significant differences in cardiac death, MACE, stroke, or revascularization. However, in the stratified analysis of RCTs, beta-blockers did not significantly improve all-cause mortality.

The lack of benefit in terms of mortality based on RCTs in the contemporary era significantly dampens beta-blockers’ role in this population, contrasting with pre-PCI recommendations. Given the variability in effect size, it is essential to consider individual patient factors when prescribing beta blockers. Personalized treatment approaches could help optimize outcomes and minimize potential adverse effects. Based upon the data available in observational studies included in our study, our findings can advocate for the continued use of beta-blockers as a standard therapeutic option in post-MI care, particularly to reduce all-cause mortality or in those at higher risk of recurrent events. However, further RCTs are required to provide results with a better strength of evidence.

A previous meta-analysis by Hu M. et al. also reported a reduced risk of all-cause mortality with beta-blocker use in patients with MI (31). However, it also showed significant results in cardiac death, myocardial infarction, and revascularization without influence on heart failure, MACE, or stroke. A meta-analysis conducted by Maqsood et al. (32) and Dahl Aarvik et al. (33) also favored the use of beta blockers in causing a reduction in all-cause mortality. However, these meta-analyses also only included data from observational studies, with Hu M. et al. having only one RCT. Our meta-analysis included 3 RCTs and propensity score-matched observational studies, thus limiting the impact of confounders on our results. Including more RCTs and observational studies with propensity score matching in our study showed differences in secondary outcomes from previous meta-analyses.

Beta-blockers have long been a cornerstone in managing patients following an MI. Our study aimed to evaluate the efficacy and safety of beta blockers in contemporary post-MI patients, considering the advances in medical therapy and revascularization techniques. The use of beta blockers in patients with acute myocardial infarction was first evidenced long before the reperfusion era when the First International Study of Infarct Trial (ISIS-1) in 1986 showed a significant reduction in vascular death with the use of beta blockers compared to non-use of beta-blockers (3.87% vs 4.57%, P-value < 0.05) (34). However, later in 2005, clopidogrel and metoprolol in the Myocardial Infarction Trial (COMMIT) showed no significant differences in all-cause death, myocardial infarction, or cardiac arrest with the use of metoprolol compared to no use of beta-blocker (P=0.01) (35). These differences in results can be attributed to the advancement in treatments in the reperfusion era. In the ISIS-1 trial, only 5% of patients received antiplatelets, whereas, in COMMIT, all patients received aspirin, with 50% receiving dual antiplatelet therapy and 54% receiving fibrinolytic agents. Advances in PCI techniques and the increased use of aspirin, clopidogrel, and statins have substantially decreased all-cause mortality. Consequently, it is hypothesized that modern treatment modalities have reduced the effectiveness of beta blockers due to advancements in the reperfusion era (36).

The potential impact of beta-blockers on patients with acute myocardial infarction (AMI) can be attributed to several mechanisms. AMI significantly increases monocyte accumulation in atherosclerotic plaques, leading to larger and more complex lesions. Additionally, the sympathetic nervous system activation activates neuroimmune connections in the bone marrow, boosting extramedullary myelopoiesis. Prior beta-blocker therapy was associated with decreased circulating monocytes after myocardial infarction, attenuating atherosclerosis and improving long-term patient outcomes (37). Adverse cardiac remodeling in the form of ventricular dilation following AMI is linked to a worse prognosis (38), and beta-blockers have been shown to improve this left ventricular remodeling process (39). Beta-blockers are class II antiarrhythmic drugs known to reduce the occurrence of both short- and long-term ventricular arrhythmias, which are major causes of 90-day mortality (40–42). Much of the benefit of beta blockers owes to the neurohormonal blockage of sympathetic drive. This results in reduced heart rate, myocardial oxygen demand, and decreased levels of circulating vasoconstrictors, leading to reduced peripheral resistance and myocardial work. All these effects are cardioprotective, helping reduce cardiomyocyte death and prevent deleterious cardiac remodeling.

A notable strength of this analysis is the inclusion of more RCTs and propensity score-matched observational studies, which enhances the generalizability of our findings to current clinical practice. Furthermore, the risk of bias assessment, detailed in Supplementary Table 1, highlights that most included studies demonstrated strong methodological quality. For instance, studies such as Choo (2013), Ishak (2023), and Konishi (2015) each scored 9 out of 9, indicating robust study designs. We also conducted an extensive literature review across multiple databases, capturing a broad spectrum of studies, stringent inclusion criteria, and robust quality assessment methods to minimize bias.

A few important limitations linked with this meta-analysis also need to be addressed. The high heterogeneity (I² = 96%) highlights the need to interpret the pooled results carefully. Factors such as differences in study design, patient demographics, and treatment protocols contribute to this variability. Additionally, the observational nature of most included studies limits our ability to infer causality. The differences in results observed between observational studies and RCTs on stratified analysis may be because our RCT-based analysis was underpowered compared to observational studies. This also brings forth the need to conduct more RCTs to develop a more accurate picture of the role of beta blockers in post-MI patients. Moreover, the RCT by Yndigegn 2024 showed a high risk of bias assessment attributed to the selection process, which might be due to an inadequate randomization process. Future RCTs are necessary to confirm these findings and address the heterogeneity observed.

## Conclusions

While our meta-analysis underscores the significant mortality benefit of beta blockers in contemporary post-MI patients in the combined analysis, it fails to confidently support the continued use of beta blockers as a crucial component of post-MI management on stratified analysis due to differential results between observational studies and RCTs. Tailoring treatment to individual patient profiles will be key to maximizing clinical benefits and improving patient outcomes

## Data Availability

All data produced in the present study are available upon reasonable request to the authors.
All data produced in the present work are contained in the manuscript as well.

## Acknowledgments

Not applicable.

## Availability of data

The data supporting this study’s findings are available from the corresponding author upon reasonable request.

## Supplementary Data

**Supplemental Figure 1:**
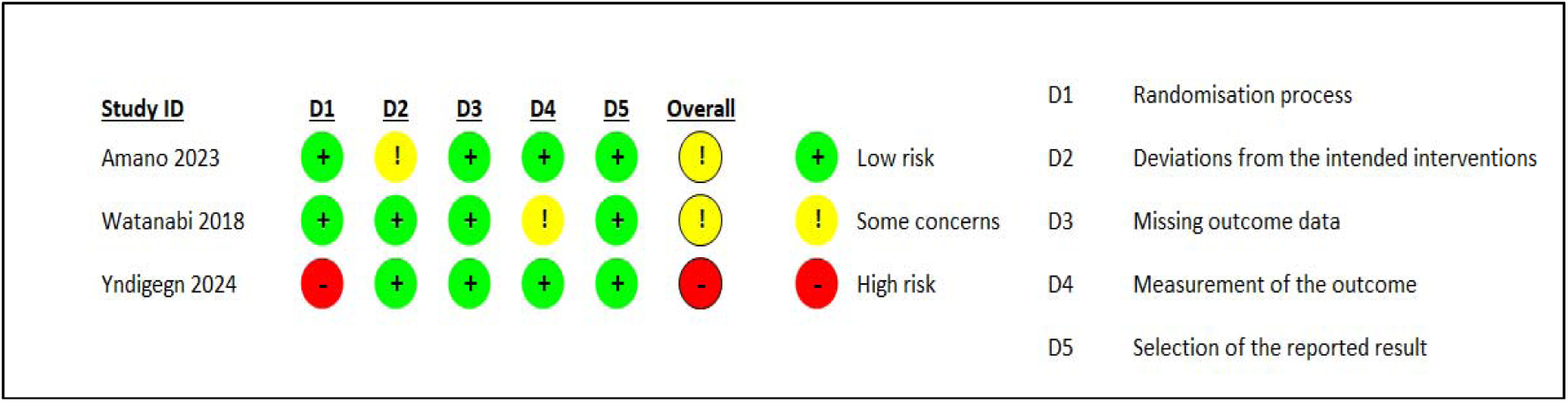
Risk of Bias in RCTs

**Supplemental Figure 2:**
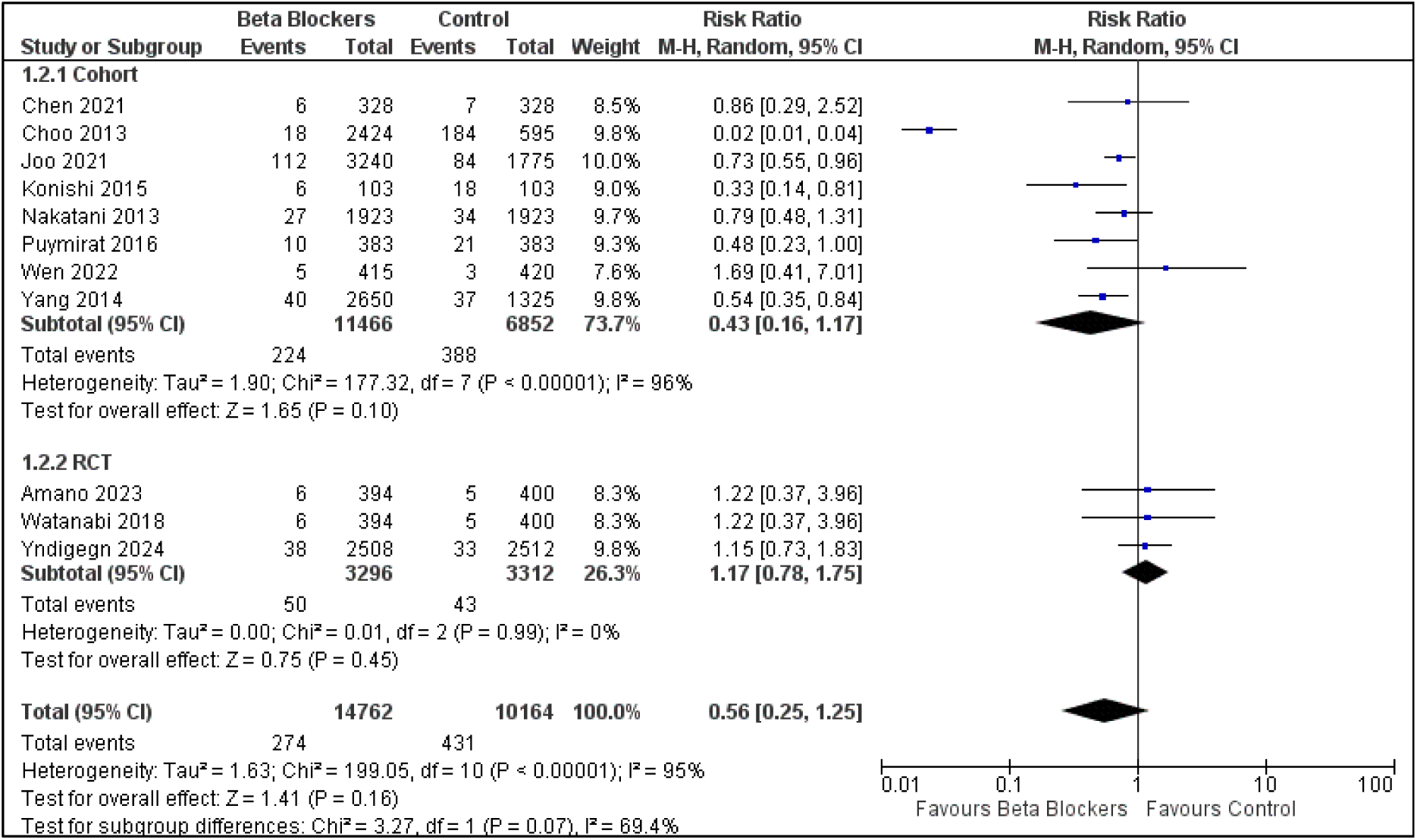
Forest Plot of Cardiac Death

**Supplemental Figure 3:**
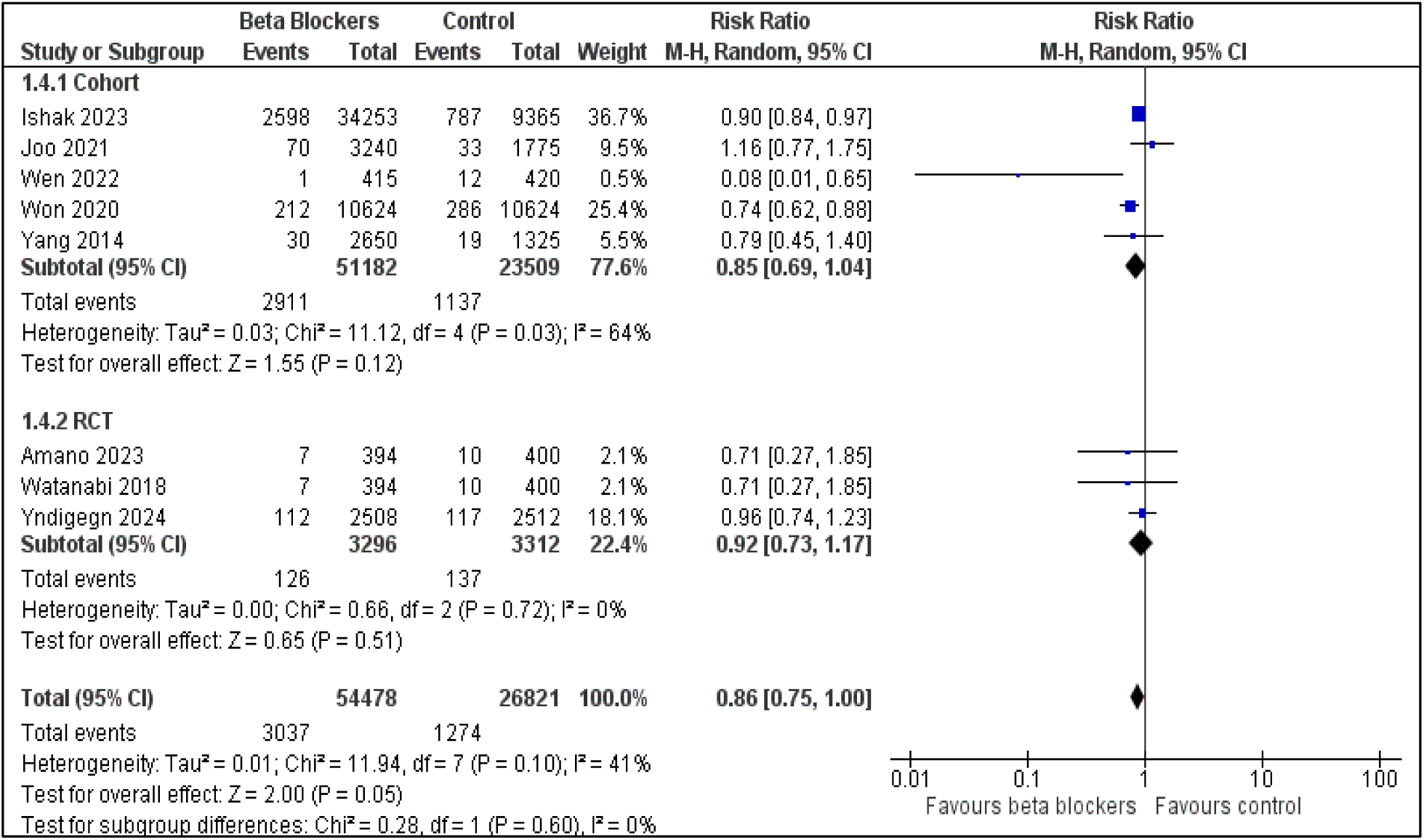
Forest Plot of Myocardial Infarction

**Supplemental Figure 4:**
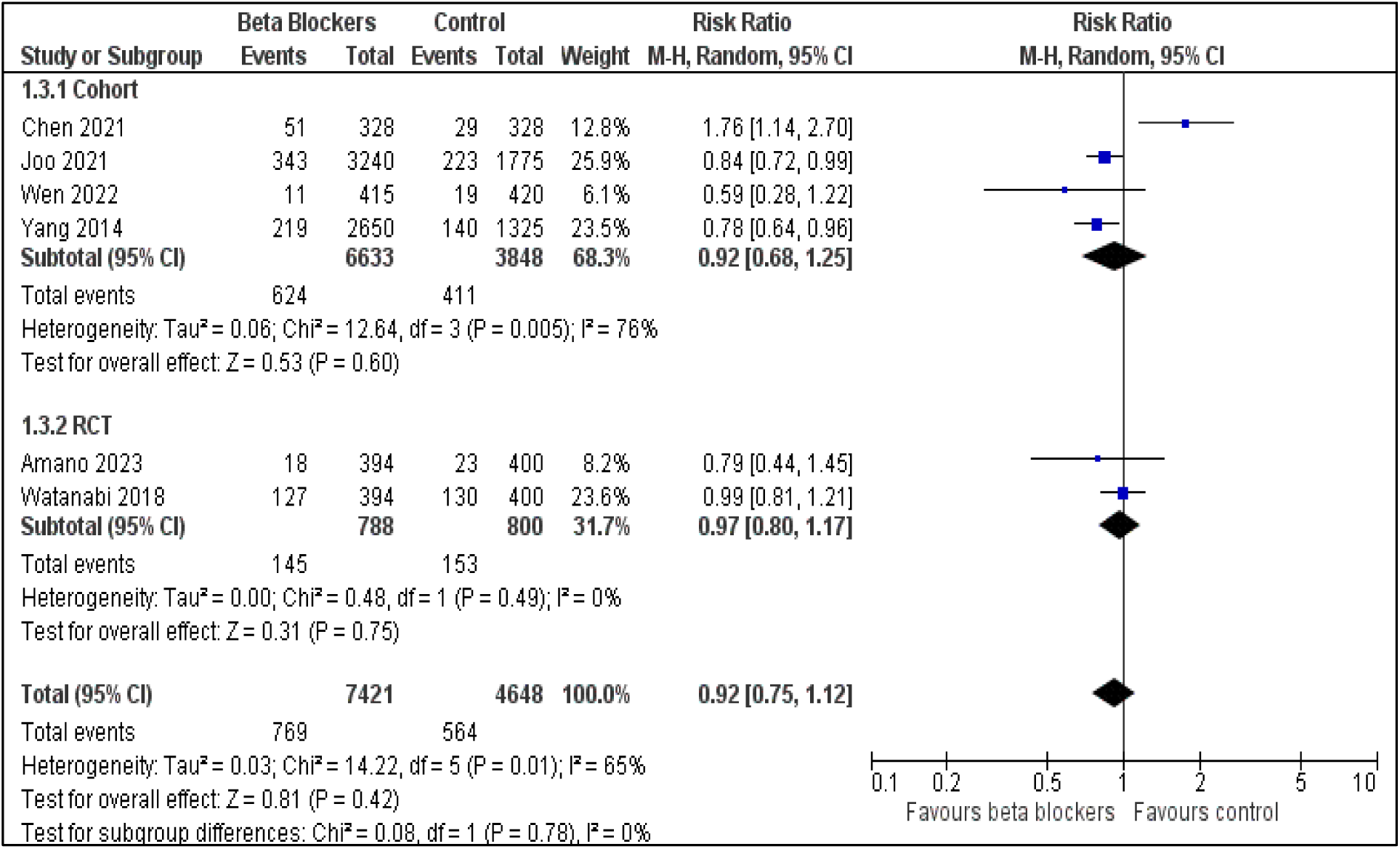
Forest Plot of Major Adverse Cardiovascular Events

**Supplemental Figure 5:**
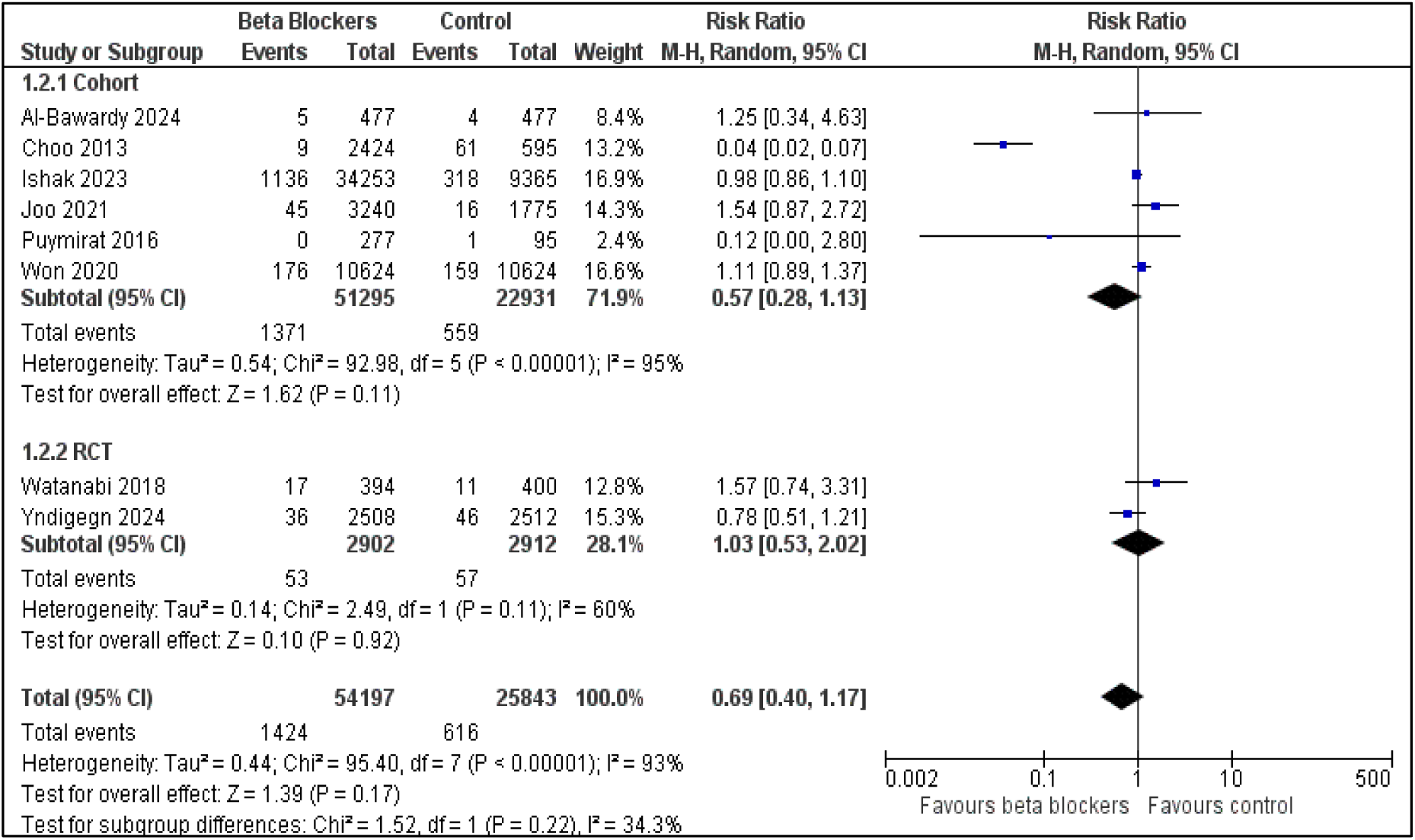
Forest Plot of Stroke

**Supplemental Figure 6:**
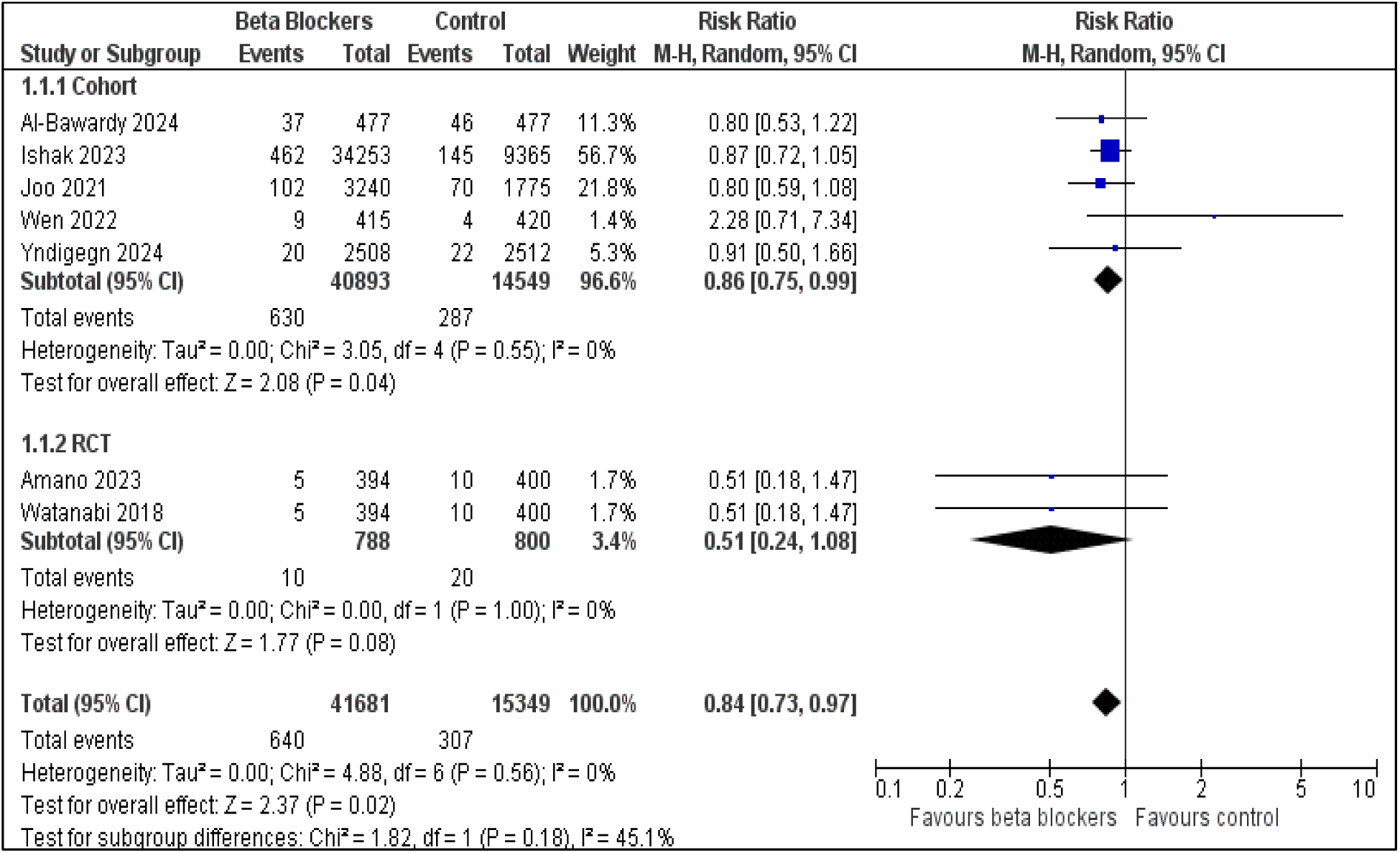
Forest Plot of Heart Failure

**Supplemental Figure 7:**
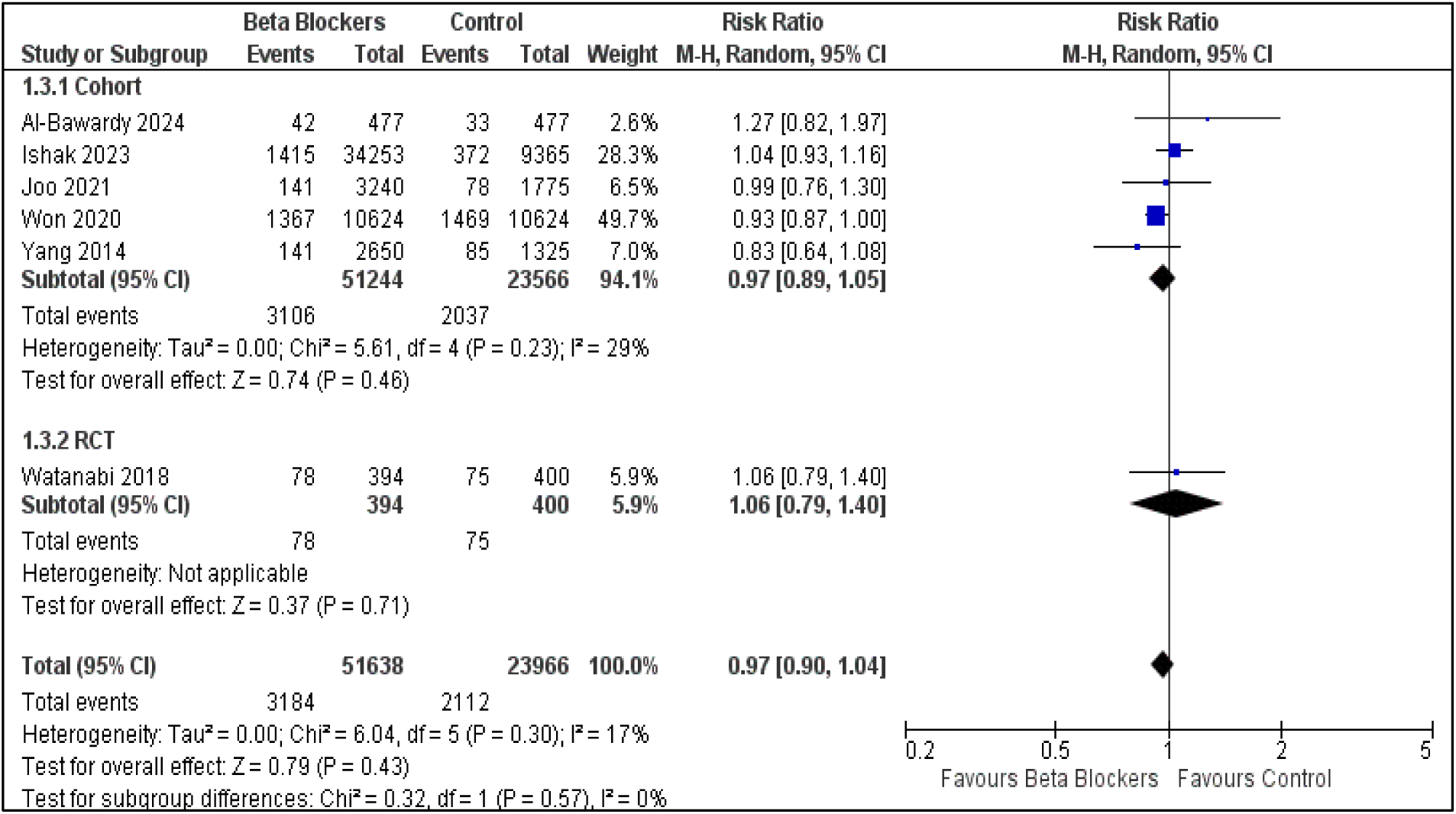
Forest Plot of Revascularization

**Supplementary Table 1:** Risk of Bias in Observational Studies.

| Study | Selection |  |  |  | Comparability |  | Outcome |  |  | Total |
| --- | --- | --- | --- | --- | --- | --- | --- | --- | --- | --- |
|  | S1 | S2 | S3 | S4 | C |  | O1 | O2 | O3 |  |
| Al-Bawardy 2016 | * | * | * | 0 | * | * | * | * | 0 | 7* |
| Chen 2021 | * | * | * | * | * | 0 | * | * | 0 | 7* |
| Choo 2013 | * | * | * | * | * | * | * | * | * | 9* |
| Ishak 2023 | * | * | * | * | * | * | * | * | * | 9* |
| Joo 2021 | * | * | * | * | * | * | 0 | * | * | 8* |
| Konishi 2015 | * | * | * | * | * | * | * | * | * | 9* |
| Lee 2020 | * | * | * | * | * | * | * | * | 0 | 8* |
| Nakatani 2013 | * | * | * | * | * | * | * | * | 0 | 8* |
| Puymirat 2016 | * | * | 0 | * | * | * | * | * | * | 8* |
| Wen 2022 | * | * | * | * | * | * | 0 | * | 0 | 7* |
| Won 2019 | * | * | * | * | * | * | * | * | * | 9* |
| Yang 2014 | * | * | * | * | * | * | * | 0 | * | 8* |

